# Professional Commitment and Its Associated Factors Among Institutional Delivery Services Providers In Public Health Facilities, In Shone District, Southern Ethiopia

**DOI:** 10.1101/2021.08.09.21261359

**Authors:** Melaku Wolde Anshebo, Tesfaye Gobeana Tessema, Yosef Haile Gebremariam

## Abstract

**Background:** There is paucity of information on level of commitment among health professionals attending delivery service in public health facilities of low-income countries including Ethiopia.

**Objective:** To assess the level and factors associated with professional commitment among institutional delivery services providers at public health facilities in Shone District, Southern Ethiopia.

**Methods:** A facility-based cross-sectional study design was conducted at primary level public health facilities in Shone District. All health facilities (one primary hospital and 7 health centers) were included in the study. 503 study participants who fulfilled inclusion criteria in proportion to obstetric care providers in each public health facilities were selected by applying simple random sampling. Self-administered Likert scale type of questionnaire was used. Data were analyzed using SPSS version 20. Bivariate and multivariable logistic regression analyses were done to see the association between dependent and explanatory variables.

**Results:** The magnitude of professional commitment for obstetric care providers working in public health facilities of Shone district was 69.4%. In this study, those who worked at hospital, those who had positive attitude toward organizational commitment, and those who had positive attitude toward personal characteristics were 2.4, 2.3 and 1.76 times more likely committed to profession compared with the counterparts respectively.

**Conclusion:** The professional commitment among institutional delivery service provision was medium. All health professional should manage their own personal characteristics to behave in good way to be committed for their profession. Organizational commitment had great influence on professional commitment.

## Introduction

Commitment is the belief in and acceptance of the goals and values of the profession and a willingness to exert considerable effort on behalf the profession and definite desire to maintain membership in the profession. Thus, any scale that purports to measure professionals’ commitment should tap one of these mind sets and should reference the target, what the employee is committed to, be it the organization, profession, a team, a change initiative, a goal which can be to the maternal health services goal (institutional delivery services) in this case (1). Meyeren and Allen stated that professional commitment has three dimensions: (a) Affective commitment, (b) Continuance commitment and (c) Normative commitments(4)

One of the most comprehensive studies of professional commitment examined not only the determinants of professional commitment but also the determinants of organizational commitment. Personal background, organizational context, and socialization variables, organizational commitment and job satisfaction can affect professional commitment (1).

Health professionals ‘commitment can be explained by adherence to principles of ethical practice, effective interactions with patients and with people who are important to those patients, effective interactions with other people working in the health care system, reliability, commitment to autonomous maintenance and continuous improvement of competence, pride in profession, appearance, flexibility and good behavior outside work(2). It is a more stable type of work commitment or occupational commitment and feelings about a job or organization (3).

Institutional delivery service is one of the essential maternal health services that is given at health facilities by trained and educated health professionals(1). Institutional delivery services utilization coverage is increasing globally including Africa, but the progress is very slow in sub-Saharan Africa including Ethiopia (5). Skilled attendance during delivery can only be provided in the presence of functioning health systems which include adequately trained, motivated and committed health workers (6).

In Ethiopia the health system is facing many challenges related to shortage of motivated and committed health professionals in different disciplines and at all levels. The factors that could affect professional commitment could be role status, salary levels, degree of autonomy over work and the level of exposure to on-going training and staff development opportunities and levels of stress in the work environments(7). Demographic characteristics such as age, gender, marital status, educational achievement and other factors like career history, work values, attitudes toward the organizational climate and personality variables could also affect professional commitment (8).

Health professional (obstetric care providers) in public health facilities is often absent during working hours and these complaints are acknowledged by health worker’s commitments. These may affect quality of maternal health services which in turn increase maternal and neonatal mortality(9).

There are limited studies conducted on the professional’s commitment and factors among obstetric care providers in Ethiopia particularly in this study settings. Thus, it is indispensable to assess the professional commitment and its related factors in different socio-economic setting of the country. Therefore, the aim of this study is to assess the level of professional commitment and its associated factors among obstetric care providers in public health facilities of Shone District, Southern, Ethiopia.

## Method and Material

### Study Area and Period

The study was conducted in Shone District, Hadiya zone, Southern Ethiopia from October 2019 to March 2020. In the district, there are one primary Hospital, seven health center, four private medium clinics and six drug stores. There are about 870 health professionals from different disciplines from these the relevant for this study were 668 who have been providing obstetric care in the public health facilities of Shone District.

### Study design

A facility based cross-sectional study design was conducted at primary level public health facilities in Shone District.

### Source Population

The source population was all health professionals who were providing institutional delivery services at public health facilities in Shone District.

### Study Population

The study population was all selected health professionals who were providing institutional delivery services at public health facilities in Shone District.

### Inclusion Criteria

All health professionals who have been employed in the health facilities for at least 6 months and directly involved in the care of pregnant women and institutional delivery service provision were included in this study.

### Exclusion Criteria

Those who were critically sick and not directly involved in institutional delivery service provision such as pharmacist, lab technician, environmental health, dentists, psychiatry nurse and radiographers were excluded.

### Sample Size Determination

The sample size was calculated using single population proportion formula as follows:

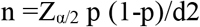

Where: n= minimal sample size of study, P =Estimated percentage of professional commitment towards institutional delivery services (p=72.71%) in study conducted in Jimma Zone Southern west Ethiopia(3), d = Marginal error/degree of precision =5% (0.05), α = Critical value at 95% CI of certainty (1.96), Z= Reliability coefficient, n =1.96^2^ (0.7271) (1-0.7271)/ (0.05)^2^ =457.

Then, by adding 10% for non-response rate, final sample size became 503.

### Sampling Techniques

Frist, the health facilities were stratified into hospital and health centers. Then, one district hospital and seven health centers were included in the study. Then, based on numbers of study participants in each facility, simple random sampling method was applied for the selection of study participants in proportion to professionals in each public health facilities.

### Data Collection Tool and Procedure

The quantitative data were collected using self-administered structured questionnaire that was adapted from different literatures based on the study objectives (3,10).

Four data collectors were involved for collecting data using quantitative self-administered questionnaire and they are HO and Nurses (BSc) holders. The participants of the pre-test were contacted to give their general feelings, comments and problems encountered while responding the questions. The study participants were invited to participate voluntarily by explaining the rationale of the study at the time of data collection.

### Operational Definition

#### Professional commitment

is the relative strength of an individual’s linkage to the respective profession. It is beyond a commitment for a particular organization and implies the individuals’ perspective towards their profession and the motivation that they have to stay in their job which refers to one’s loyalty to the profession and the willingness to strive and uphold the value and goals of the profession to maintain membership in the profession that can be explained by adherence to principles of ethical practice, effective interactions with patients and with other people working in the healthcare system, reliability, commitment to autonomous maintenance and continuous improvement of competence, pride in profession & good behavior outside work. This were measured using 10 items of 5-point Likert scale and 1 denoting for commitment, 0 denoting for uncommitted. Professional commitment score was created and higher score indicates higher professional commitment. The mean scores for all scales was reported as the percentages of scale mean score (%SM) after standardization the mean was calculated(3,11).

### Data Quality Control

The questionnaire was pretested on 5% of the actual sample size at Balesa health center in Lemo district before the actual data collection period to make correction on tool accordingly. Training was given for both data collectors and supervisors for one day by the investigator. There was supervision on daily basis. Finally, verification was done by checking error report before entry to Epi data using each case code. Split sample validation and outliers ‘detection done.

### Data Processing and Analysis

Professional commitment, which is the outcome variable was measured using ten questions, the index value was generated using descriptive statistics, such as means, summary tables, and graphs was used for describing the data. First, data was checked manually by principal investigator for its completeness and consistency. Each completed questionnaire was assigned a unique code and entered to EpiData version 3.02 Software. Then, data were exported to SPSS version 20.0 for analysis. Multi-collinearity was checked by examining the variance inflation factors (VIF), so if the values for each variable is less than or equal to 5 was taken as no similarity or correlation coefficient <0.9. Hosmer and Lemeshow goodness test were used to check model fitness. Bivariate analyses were used to assess the relationship between dependent and independent variables. Those variables with p-value of<0.25 in bivariate logistic regression were inserted into the final model. Professional commitment mean scores between health facilities (HC and hospitals) professionals were computed after checking the assumption that whether the difference in mean commitment level between different group was statically significant or not. Factors associated with professional commitment were identified using multivariable logistic regression analysis at a significance level of p-value < 0.05 with 95% confidence interval.

### Ethical consideration

The study was approved by Haramaya University, College of Health and Medical Sciences Institutional Health Research Ethics Review Committee (IHRERC). The permission agreement consent was taken from south nation nationality people region health bureau, Shone district health department and respective woreda health offices. Informed, Voluntary, Written & Signed consent was obtained from the Heads of health facilities. Participants was informed clearly about the purpose and benefit of the study, and Informed, Voluntary, written and signed consent was obtained. Those who signed written consent were only participated in the study and the confidentiality of responses was maintained throughout the research process by giving code for participant. They were informed well that they have full right to totally refuse to participate and/ or draw from the interview at any time of they have any problem.

## Results

### Socio-demographic characteristics of study participant

Of the total of 503, 480 (95.42%) were participated in the study. More than half (50.6%, n=243) of the respondent were males. Similarly, more than half of the respondents were married (53.3%, n=256). Most of the respondents (71.7%) were in the age category of 24-34 years (table 1). Almost half (49.2%) of the respondents were nurses in their profession. Once more, more than half (54.2) of respondents had work experience of 2-5 years in labor ward. Nearly two third (65.6) of the respondents had work experience of 2-5 years in the health facility. Of the total of 480 respondents; 162 (33.8%), 200 (41.7%) and 118 (24.6%) were dwellers of rural, urban and semi-urban areas, respectively. Moreover, about two third (66.3%) of the respondents were workers in health centers. Furthermore, nearly half (47.1) of the respondents had monthly salary in the range from 3654 Birr to 5294 Ethiopia Birr (Table 1)

**Table 1:**
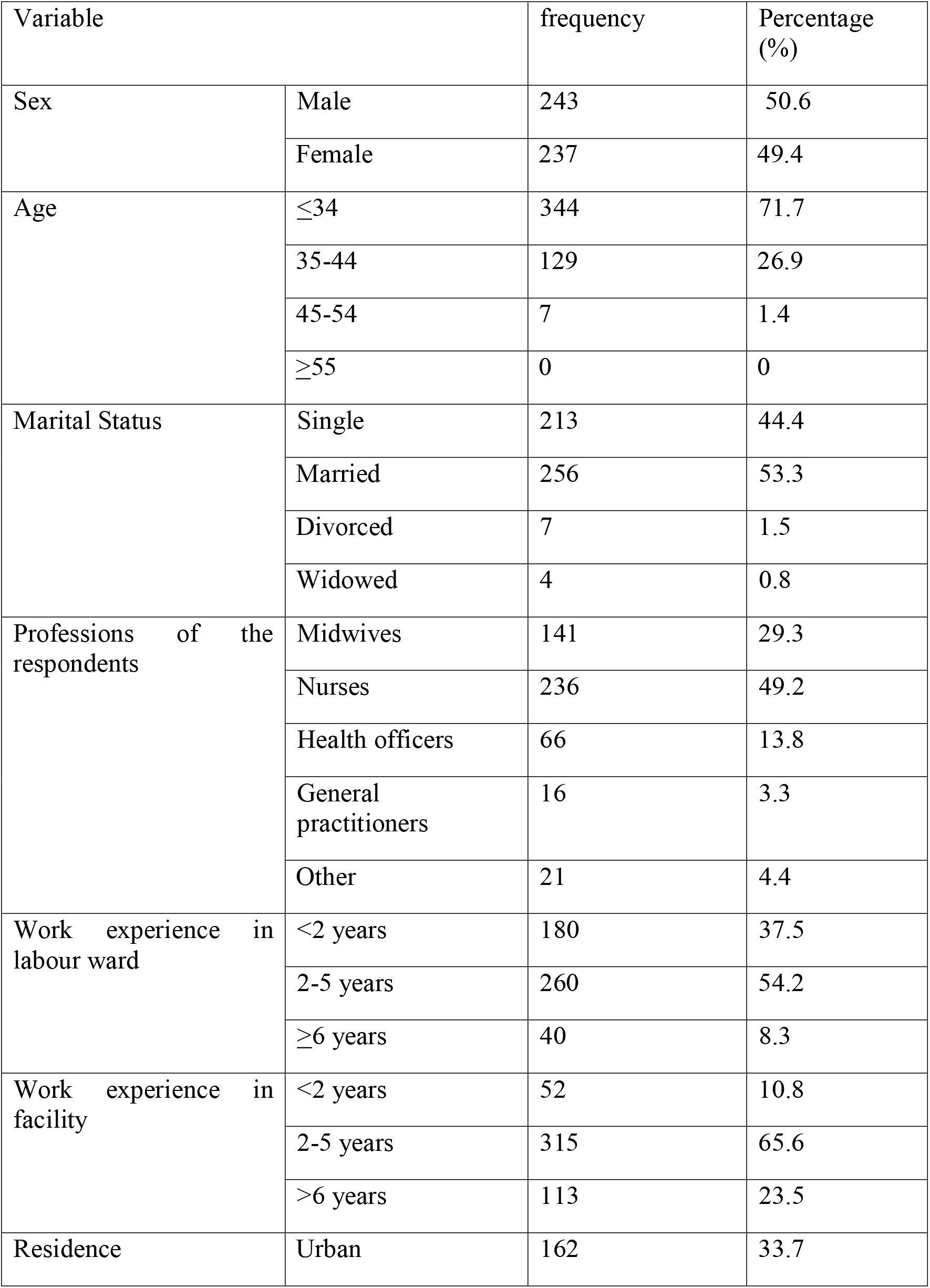
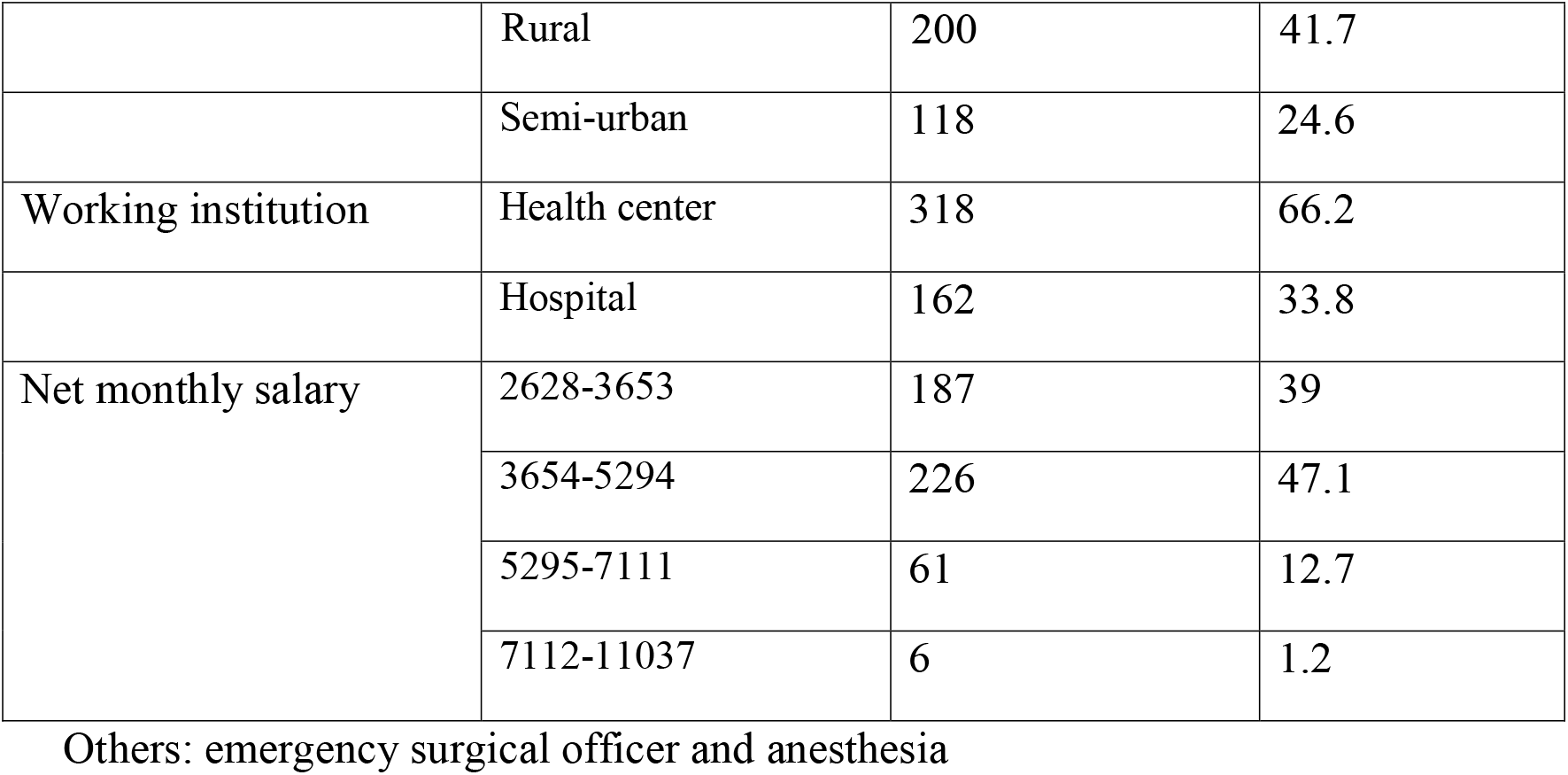
Socio-demographic characteristics of health professionals, Shone district, Hadiya Zone, Southern Ethiopia, from October 2019 to June 2020 (n=480).

### Level of Professional Commitments

Out of 480 study participants; about half and 21.3% of them agree and strongly agree the idea that they had willing to put a great deal of effort to develop their profession respectively. Similarly, about 46.5% and 30% of the respondents agree and strongly agree that they were identified by their profession respectively. Likewise, nearly half (48.5%) and more than one third (33.8%) of the participants agree and strongly agree the perception that they would accept any type of job related to their profession to reduce complaints accompanied with it respectively. Once more, about 46% and 37.1% of the respondents agree and strongly agree the idea that they felt strongly tied with their colleagues respectively. Moreover, about 45.4% and 27.7% of the study participants agree and strongly agree the perception that they were proud to belong to their profession, respectively. Furthermore, nearly half (48.5%) and one third (29.8%) of the respondents agree and strongly agree the idea that they really inspired in their best in the way of job performance, respectively (Table 2).

**Table 2:** Frequency distribution of five-point Likert scale response of participants for professional commitment measuring items, Shone district, 2020 (n=480).

| Items measuring professional Commitment. | Strongly disagree | Dis agree | Neutral | Agree agree | Strongly |
| --- | --- | --- | --- | --- | --- |
| I am willing to put in a great deal of effort to develop my profession beyond expected. | 9(1.9%) | 81(16.9%) | 46(9.6%) | 242(50.4%) | 102(21.3%) |
| I am a person who identifies strongly with my profession. | 7(1.5%) | 63(13.1%) | 43(9.0%) | 223(46.5%) | 144(30.0%) |
| I would accept almost any type of job that related to my profession to keep working beyond expected from me. | 4(0.8%) | 47(9.8%) | 34(7.1%) | 233(48.5%) | 162(33.8%) |
| I am a person who feels strong ties with other members of my profession. | 2(0.4%) | 54(11.3%) | 24(5.8%) | 222(46.0%) | 178(37.1%) |
| I am a person who is proud to belong to my Profession. | 0 | 64(13.3) | 65(13.5%) | 218(45.4%) | 133(27.7%) |
| My profession really inspires the very best in me in the way of job performance. | 2(0.4%) | 67(14.0%) | 35(7.3%) | 233(48.5%) | 143(29.8%) |
| I am extremely glad that I chose this profession to work for ever in advance. | 5(1.0%) | 54(11.3%) | 23(4.8%) | 255(53.1%) | 143(29.8%) |
| I am a person who criticizes my profession. | 36(7.5.9%) | 121(25.2%) | 39(8.1%) | 181(37.7%) | 103(21.5%) |
| I am considering my profession to be important | 5(1.0%) | 63(13.1%) | 25(5.2%) | 206(42.9%) | 81(37.7%) |
| I am a person who tries to hide belonging to my profession. | 132(27.5%) | 168(35.0%) | 34(7.1%) | 100(20.8%) | 46(9.6%) |

### Magnitude of professional commitment

The magnitude of professional Commitment was 69.4%, 95% CI (65, 74) of health professionals. More than half 53.1% and nearly one third 29.8% of the respondents agree and strongly agree the idea that they were extremely glad to choose their profession to work for life in advance respectively (Figure1)

**Figure1:**
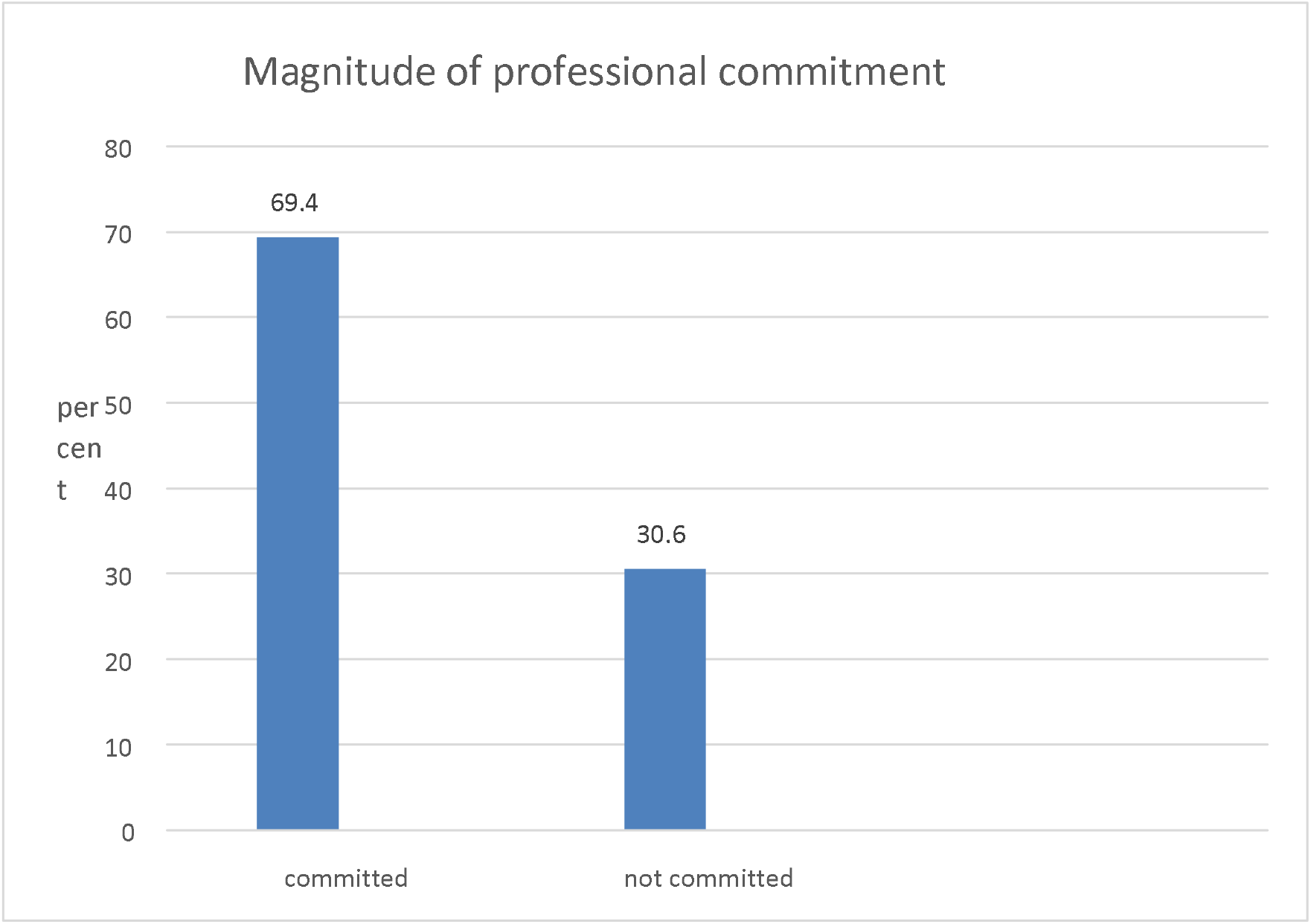
Magnitude of professional commitment in government health facilities in Shone district, Southern Ethiopia, from October 2019 to June 2020.

### Factors associated with professional commitment

In binary logistic regression analysis, profession of the respondents, work experience in labour ward, area of residence, type of health facility, net monthly salary, job satisfaction, organizational support, organizational commitment, personal characteristics and institutional delivery service were found to be associated with professional commitment. These 10 variables, which had statistically associated with professional commitment in the preceding in binary logistic regression model were entered into the final model (Multivariable logistic regression model). Out of that only three of them were significantly associated with professional commitment. (Table 3)

**Table 3:** Multivariable analysis of factors related with professional commitment in public health facilities, Shone district, Hadiya Zone, Ethiopia, 2020 (n=480)

| Variables | Professional commitment |  | COR<br>(95%CI) | AOR |
| --- | --- | --- | --- | --- |
|  | Committed | Not<br>committed |  |  |
| Types of health facility |  |  |  |  |
| Health centers | 205(65.5%) | 113(35.5%) | 2.06(1.33-3.22) | 2.45(1.29-4.67) ** |
| Hospital | 128(79.0%) | 34(21.0%) | 1 | 1 |
| Job Satisfaction |  |  |  |  |
| Dissatisfaction | 197(64.0%) | 111(36.0%) | 1 | 1 |
| Satisfaction | 136(40.8%) | 36(20.9%) | 2.13(1.38-3.29) | 1.72(0.72 – 2.47) * |
| Organizational Support |  |  |  |  |
| Negative Attitude | 191(64.5%) | 105(35.5%) | 1 | 1 |
| Positive Attitude | 142(77.2%) | 42(22.8%) | 1.86(1.22-2.82) | 1.03(0.571 -1.856) * |
| Organizational commitment |  |  |  |  |
| Negative attitude | 128(59.0%) | 89(41.0%) | 1 | 1 |
| Positive attitude | 205(77.9%) | 58(22.1%) | 2.45(1.65-3.65) | 2.27(1.25-4.12) ** |
| Personal characteristics |  |  |  |  |
| Negative attitude | 86(53.4%) | 75(46.6%) | 1 | 1 |
| Positive attitude | 247(77.4%) | 72(22.6%) | 2.99(1.99-4.49) | 1.76(1.04-2.96) ** |

AORs with CI 95% indicated with bold shows variables which associated with outcome variable with p- value < 0.05\*=p>0.05, \*\*=p<0.05, \*\*\*=p<0.001

A multivariable logistic regression analysis was performed to identify factors associated with professional commitment of health care professionals. Consequently, three variables were found to be statistically associated with professional commitment after adjusting for confounders. These were type of facility, organizational commitment and personal characteristics. In this study, those who worked at hospital were 2.4 times more likely committed to profession compared with those who worked at health center [AOR=2.45:95%CI (1.29-4.67)]. Those who had positive attitude toward organizational commitments were 2.3 times more likely committed to profession compared with those who had negative attitude toward organizational commitment [AOR=2.27:95% CI (1.254.12)]. Those who had positive attitude toward personal characteristics were 1.76 times more intended to professional commitment compared with those who had negative attitude toward personal characteristics [AOR=1.76:95%CI (1.04-2.96) p=0.03] (Table 4).

## Discussion

This study finding shows the level of professional commitment was 69.4%. The finding is lower than other study findings done in Ghana and Jimma, south eastern Ethiopia which revealed that level of professional commitments were 73% and 72.7%, respectively(3,12). The possible explanation for the disparity might be due to variation in working environment and the study setting as well. This study finding is higher than the study result done in Gurage Zone, Southern Ethiopia which showed that the proportion of professional commitment to their organization was 64.81%(11). The possible explanation might be due to improvement of promotion opportunities and other incentives like housing loan, medical benefit (risk loan, the way of pay of duty) which were provided by the organization to the professionals.

The other study finding done in India, showed that good relation with organization, fair training and development policy, clear promotion policy, cost to live on the same organization could help in delivery responsibilities’ professionals that may add the professional’s commitment (13).

The other finding of this study showed that health professionals working in health center were 2.45 times more likely to be committed to their work than professionals working in hospital. This finding is consistent with study findings done by Havid (14). The reason for the difference of professional commitment between health professionals working in hospital and health centers might be due to decreased work overload in health centers than hospitals.

Similarly, this study result reveals that health professionals who had positive attitude to their organization were 2.27 times more likely to be committed to their professions compared to health professionals who had negative attitude to their organization. This finding is in agreement with other study finding done in USA, New York which stated that Physicians loyal to their organization were committed to their professions.

Health professionals who had positive attitude to personal characteristics were 1.76 times more likely to be committed to their work compared with health professionals who had negative attitude to personal characteristics. This finding is also consistent with other study findings done in New York, USA which stated that belief in individual physician autonomy and belief in collective self-regulation were associated with professional commitment positively.

Consequently, three variables were found to be statistically associated with professional commitment after adjusting for confounders. These were types of health facility, organizational commitment and personal characteristics. Organizational commitment related predictors of professional commitment were organizational affective (affection for job) commitment factors and organizational normative (Sense of obligation to stay) commitment factors. Related researches support this finding by phrasing the affective component (AC) is adoption of organizational goals and commitment to them and to have positive emotions related to identification with it.

Likewise, this study result agreed with other study done in Bangladesh, which showed that work environment had significant, moderate, and positive relationship to their organizational commitment(15). This study finding shows the personal characteristic, which is nearly half of respondent were work with absenteeism which is higher than in study done Israel (16). The finding of this study also showed that health professionals who were always on time for work was higher than the study done in the Norway, which showed that they were always on time for work was reported by individuals was 4.315 in 1993 and 4.293 in 2003, and the correlation (r=0.444) (17).

## Conclusion

The magnitude of professional commitment among institutional delivery service providers in public health facilities in Shone district was medium. Personal characteristics was an important factor of professional commitment. More specifically, occupational stresses and having balanced work life time to foster a higher level of professional commitment. Organizational commitment had great influence on professional commitment. Specifically, affective and normative organizational commitment score affect the level of professional commitment positively. Moreover, the perceived work environment and perceived team work of the staff were to increases professional commitment. Finally, we can conclude that professional commitment was much more influenced by organizational related factors than personal factors by contributing many variables that can affect outcome of interest.

## Limitation

The limitations of the study were information bias and social desirability bias as well as the study couldn’t identified cause and effect relationship. The tool was subjected to information contamination by attitude of coworkers that was controlled by informing participants to complete privately.

## Data Availability

All documentary data and literature relevant to the study are publicly available. Interview data will not be shared to maintain confidentiality

## Acknowledgment

First, we would like to extend our gratitude for Haramaya University, for technical support to conduct this research. Second, we would also like to extend our appreciation to my advisors: Dr. Tesfaye Gobeana and Dr. Nega Assefa. We also acknowledge data collectors, supervisors and participants for their respective contribution.

## Contribution of Authors

MW had carried out the overall design and execution of the study, performed data collection, and statistical analysis and TG, NA and YH have conceived the study, critically revised the design of the study, data collection techniques, helped the statistical techniques and drafted the manuscript. All authors read this manuscript and finally approved for submission.

## Disclaimer

The funders had no role in study design, data collection and analysis, decision to publish or preparation of the manuscript.

## REFERENCES

1. Khanifar H, Jandaghi G, Shojaie S. Organizational consideration between spirituality and professional commitment. Eur J Soc Sci. 2010;12(4):558–71.

2. Elias RZ. The impact of professional commitment and anticipatory socialization on accounting students’ ethical orientation. J Bus Ethics. 2006;68(1):83–90.

3. Siraneh Y, Ololo S, Tsega G, Yitbarek K, Adamu A, Erchafo B, et al. Level and Factors Associated with Professional Commitment of Health Professionals Providing Institutional Delivery Services in Public Health Facilities, Southwest Ethiopia. Ethiop J Health Sci. 2018;28(4):495–504.

4. Meyer JP, Allen N. Commitment to Organizations and Occupations□: Extension and Test of a Three-Component Conceptualization. J Appl Psychol. 1993;(December 2017):1414–26.

5. Bogale D and, Markos D. Institutional Delivery Service Utilization and Associated Factors among Child Institutional delivery service utilization and associated factors among child bearing age women in Goba Woreda , Ethiopia. J Gynecol Obstet. 2014;(October).

6. Admassie, Assefa, Ali SN, May JF, Megquier S, Moreland S. “The Demographic Dividend: An Opportunity for Ethiopia’s Transformation,” Association. 2015;(October):1–43.

7. Giffords ED. The Internet and social work: The next generation. Fam Soc. 2009;90(4):413–8.

8. Chaulagain H, Rodrigues H, Spacone E, Varum H. Seismic response of current RC buildings in Kathmandu valley. Struct Eng Mech. 2015;53(4):791–818.

9. Dagne T, Beyene W, Berhanu N. Motivation and Factors Affecting It among Health Professionals in the Public Hospitals, Central Ethiopia. Ethiop J Health Sci. 2015;25(3):231–42.

10. Bishwajit M, Khumyu A, Boonyanurak P. Relationships between organizational commitments, supervisory support and job satisfaction of nurses in a public specialized hospital, Bangladesh. Bangladesh J Med Sci. 2016;15(1):39–43.

11. Hailemicael Nima G. Organizational Commitment of Health Professionals and Associated Factors in Government Health Facilities of Gurage Zone, South Ethiopia. Clin Med Res. 2016;5(5):82.

12. Honyenuga BQ, Adzoyi PN. PROFESSIONAL COMMITMENT OF NURSES IN GHANA□: THE CASE OF THE VOLTA REGION. Asia Pacific J Bus Manag. 2012;3(1):12–27.

13. Maheshwari S, Bhat R, Saha S. Commitment among state health officials & its implications for health sector reform: Lessons from Gujarat. Indian J Med Res. 2008;127(2):148–53.

14. Havig AK, Skogstad A, Veenstra M, Romøren TI. The effects of leadership and ward factors on job satisfaction in nursing homes: A multilevel approach. J Clin Nurs. 2011;20(23–24):3532–42.

15. Kumar D, Hossain Z, Jebin A. Organizational Commitment among the Employees of NGOs in Bangladesh□: An Organizational Commitment among the Employees of NGOs in Bangladesh□: An Empirical Study. Am J Trade Policy. 2019;(December 2016).

16. Cohen A, Golan R. Predicting absenteeism and turnover intentions by past absenteeism and work attitudes: An empirical examination of female employees in long term nursing care facilities. Career Dev Int. 2007;12(5):416–32.

17. Mutasim N, Rahman A, Hanafiah MH. Commitment to organization versus commitment to profession□: conflict or compatibility□? 2002;21:77–94.

